# An Open Labelled, Active Controlled, Three-Arm, Parallel-Group study of the safety and efficacy of the oral formulation of Oral Iodine Complex (RENESSANS) administered alone and in combination with standard interferon therapy in patients suffering from Chronic HCV Hepatitis

**DOI:** 10.1101/2020.06.27.20141473

**Authors:** Ghiasun Nabi, Muhammad Nasir, Ghias ulhasan, Israr Toor, Fawad Zia, Imran Hassan, Muhammad Faisal Nadeem

## Abstract

Iodine has strong antimicrobial properties and has been used in topical applications as antiseptic. Its systemic use in form of iodine complexes derived from dried seaweed extract is beneficial in treating various disorders. Hepatitis C Virus (HCV) chronic infection is present in 6-10% of the Pakistani population and is a major healthcare burden that could benefit from innovative therapeutic regimens.

**Objective:** A pilot study has shown the safety and efficacy of iodine complexes in chronic Hepatitis C virus infection. and this clinical study is aimed to further explore the previous findings.

**Methods:** This is an open-labeled, active-controlled, three-arm, parallel-group study including 90 patients of chronic HCV infection with each arm having 30 patients. The patient groups received 15mg/day iodine complex only, the standard of care therapy interferon+ribavirin, and iodine complex in combination with interferon+ribavirin regimen for 6 months. Efficacy assessment will base upon post-treatment Rapid Virological Response (RVR) at 4 weeks, Early Virological Response (EVR) at 12 weeks, and End of Treatment Viral Response (ETR) at 24/48 weeks.

**Results:** As only 3.33% of patients showed at the ETR with iodine complex alone, combination with interferon+ribavirin showed significant improvement in comparison to interferon+ribavirin alone. Iodine complex+ interferon+ribavirin showed 80% RVR and 90% ETR while the standard of care therapy showed 66.7% RVR and 76.67% ETR, respectively. No additional adverse events of iodine complex were observed.

**Conclusion:** Iodine complex showed a significant synergistic effect when combined interferon+ribavirin regimen and could be useful in relapsers and non-responders.

## INTRODUCTION

Chronic Hepatitis C Virus (HCV) infection is one of the common causes of chronic liver disease (1). Approximately 6-10% of the general public in Pakistan is expected to be suffering from this infection (2,3). Approximately, 130 – 150 million people worldwide are infected with the Hepatitis C virus (HCV) which poses a significant health care burden (4). In case that these patients are not treated at an appropriate time course, they may develop the sequelae of the chronic liver disease e.g. cirrhosis of the liver, Ascites, and Hepatocellular carcinoma (5). These factors, as well as suboptimal response rates to standard therapy, have led to an extensive search for novel therapy (6).

In the past, quite a few natural products have been tested to assess their hepatoprotective activity and possibly antiviral activity as well. These include Vitamin C (Ascorbic acid), Vitamin E, Zinc, Silymarin, Red beetroots, crushed licorice, artichoke, and the list goes on(7–9).

Iodine, Potassium iodide, and Ascorbic acid are natural products used in the management of Thyroiditis (10,11) and local surgical prophylaxis (12). Moreover, these agents have been tested in the treatment of oligomenorrhea and ovarian polycystic fibrosis, too (13,14). Iodine is one of the most effective antiseptics and surgical disinfectants for decades now and possesses great virucidal, bactericidal, and fungicidal properties (12). As virucidal, iodine is known to kill enveloped and non-enveloped viruses quite effectively (15).

A dried seaweed, kelp, is the richest natural source of complexed iodine and also contains vitamins, minerals. It has traditionally been used for many ailments including weight loss and as a galactagogue (16). Oral complex iodine compounds could be beneficial in treating many ailments, especially in the organs which take up iodine actively.

Previously, a feasibility study in which oral Iodine Compound {RENESSAN} was given to patients suffering HCV related Chronic Active Hepatitis and anti-viral activity and safety has been analyzed. In that study, RENESSAN containing regimen has been well tolerated by all the patients and has shown some antiviral activity (17).

The earlier described clinical data prompted the current study, whose objective was to assess the safety and antiviral efficacy of RENESSANS alone or in combination with conventional Interferon and Ribavirin for the treatment of the patients with chronic Hepatitis C.

## MATERIAL AND METHOD

### SELECTION OF PATIENT

Eligible patients were previously untreated adults of 18-65 yrs of age who had chronic hepatitis C based on the presence of anti HCV or detectable serum HCV RNA. All patient were excluded if they previously failed to respond to interferon-based therapy, were unable to take oral medication, unable to use contraception, had other cause of liver disease, were co-infected with HBV, had h/o alcoholism or had other concomitant medical condition like hemoglobinopathies or poorly controlled Diabetes Mellitus that would make it unlikely that the treatment could be completed.

### STUDY DESIGN

This open-labeled, active-controlled, three-arm parallel-group study was performed at medical unit 1, Lahore General Hospital/PGMI Lahore. The study protocol was approved by the institutional ethics committee. A total of 90 patients were enrolled in the study and written informed consent was signed by all the patients. This trial was registered at clinicaltrial.gov (registration no. NCT01463592) (18).

The sample size of 30 patients in each group was calculated prospectively to provide greater than 80 % power to detect a 35 % difference in proportional response rate using the Fisher’s exact test with 2 sided significance level of 0.05. all the eligible patients were randomized sequentially in one of the treatment groups. Lab personnel were blinded as to the treatment group assignment. Participants and investigators were not blinded. Side effects were assessed according to standard protocol (clinical trials group toxicity grading scale) and managed according to international guidelines.

### ASSESSMENT OF EFFICACY

The primary efficacy parameter was SVR 24 weeks after the end of treatment. Secondary efficacy of parameters included rates of rapid virological response (RVR), defined as serum HCV RNA below the limits of detection after 4 weeks, Early Virological Response (EVR), defined as serum HCV RNA below the limits of detection after 12 weeks and End of treatment response (ETR), defined as serum HCV RNA below the limits of detection after the end of treatment (24/48) weeks. Also, the rates of ALT normalization were assessed at multiple time points. Safety parameters that were assessed included adverse events and laboratory safety tests, which were performed at each visit.

### STATISTICAL ANALYSIS

The primary efficacy analysis was set to compare the proportional SVR rates for patients in the Renessans plus interferon plus Ribavirin group and interferon plus Ribavirin group using a 2-sided Fisher’s exact test with an α value of .05. Proportional interim virological response rates were compared using the chi-square or Fisher’s exact tests. Mean reductions in serum HCV RNA levels from baseline to week 4 of combination therapy were compared using analysis of variance and the Student *t-*test. Reductions in serum HCV RNA during the 12-week lead-in phase were analyzed using repeated-measures analysis of variance. All efficacy analyses were conducted on an intention-to-treat basis and dropouts were treated as failures. Exploratory logistic regression analyses were conducted to identify demographic or disease-related characteristics predictive of an SVR. Analyses were performed using SPSS statistical software version 16.

## RESULTS

### PATIENT CHARACTERISTICS

A total of 90 treatment-naïve patients were enrolled and the study was completed, including the follow-up period, in September 2013. All safety and efficacy analyses were based on the 90 patients who received at least 1 dose of medication in Fig.2. Baseline host and viral characteristics were similar among the three groups in Table 1.

**Fig. 1:**
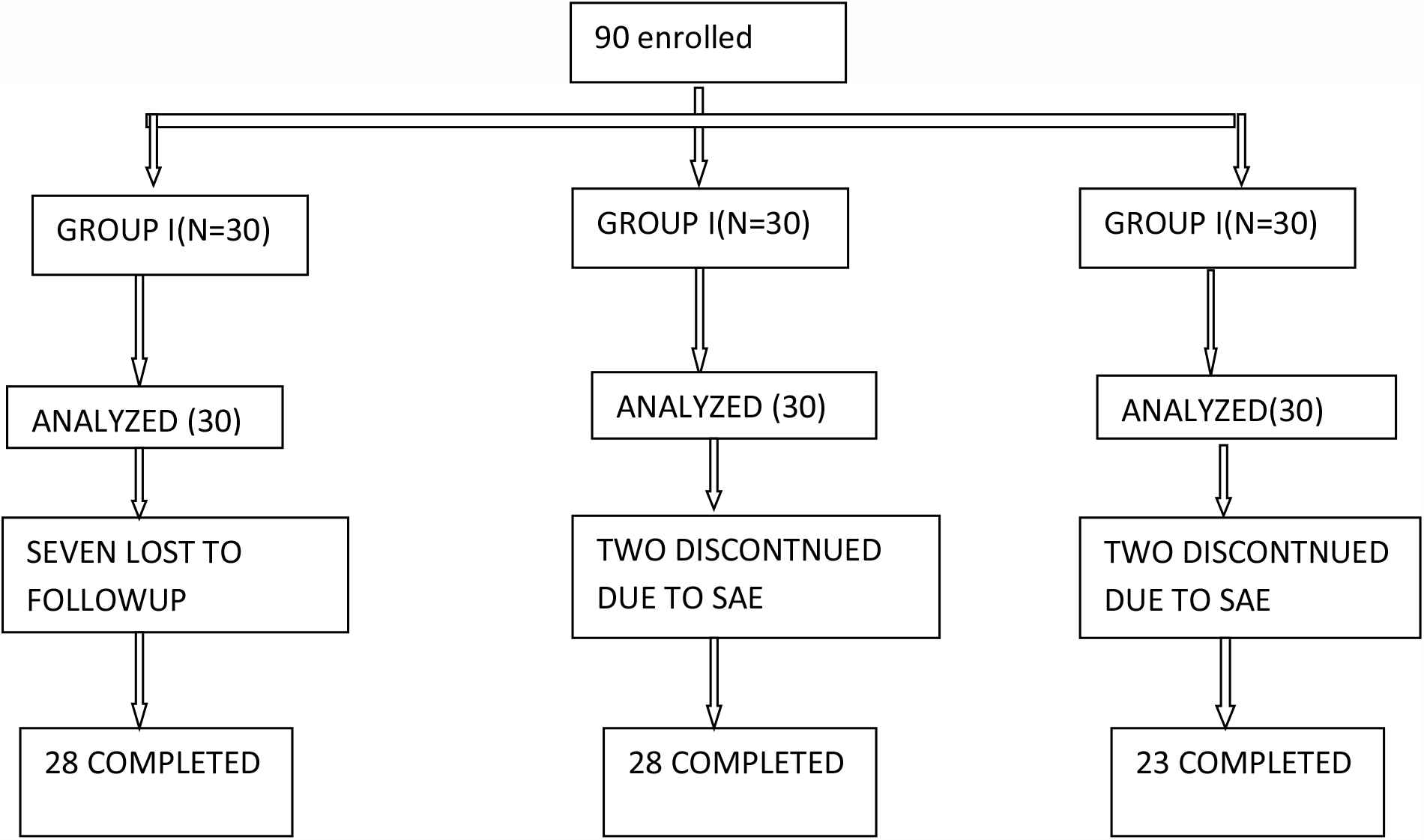
Diagram of allocation of HCV infected patients to different groups.

**Table 1:** Patients were randomized into three treatment groups.

| Study Arm | Dosing Schema |
| --- | --- |
| Group I [<br>N=30] | ORAL RENESSANS 5gm. Thrice Daily{ active iodine 300mg } for 6months |
| Group IIa<br>[N=30] | ORAL RENESSANS 5gm. Twice daily +INTERFERON ALPHA 2b 3 Million Units s/c +RIBAVIRIN 15mg/kg |
| Group II b<br>[N=30] | INTERFERON ALPHA 2b 3 Million Units s/c +RIBAVIRIN 15mg/kg |

Patients in group 1 received oral Renessans (5gm thrice daily) for 6months, Group II a received triple therapy ( ORAL RENESSANS 5gm thrice daily +Interferon Alpha 2b 3 Million Units s/c +Ribavirin 15mg/kg) and Group II b received dual therapy (Interferon Alpha 2b 3 Million Units s/c +Ribavirin 15mg/kg), patients in group II(a+b) had received response guided therapy i-e patients achieving RVR and EVR were treated for 6 months and 12 months respectively.

### RESPONSE

SVR and virological response at different time points are shown in Table 2. More patients receiving triple therapy experienced SVR compared to standard of care (86.6% vs 73%, P=0.021). Interim virological data also are displayed in table 2. The RVR rate with triple therapy compared with standard of care was higher (80% vs 66.7%, p=0.02), similarly, EVR and ETR rates are also higher in triple therapy compared with standard of care.

**Table 2.** Demographic and Disease-Related Characteristics of Patients.

|  | Group I<br>REN only<br>n=30 | Group II a<br>REN+IFN+RIB<br>n=30 | Group II b<br>IFN+RIB<br>n=30 |
| --- | --- | --- | --- |
| Age | 40± 10 | 39±8 | 38±7 |
| Median age | 39.0(20-57) | 38.5(21-55) | 38.5(23-53) |
| Sex, male/female | 15/15 | 13/17 | 14/16 |
| BMI | 27±5 | 28±4 | 26±4 |
| median BMI ( range) | 27 (18-36) | 28(20-38) | 26 (18-33) |
| Baseline HCV RNA level (log 10 IU/ml) | 5.9±0.5 | 5.8±0.4 | 5.8±0.4 |
| Median HCV RNA level( log 10 IU/ml) | 5.7 (4.2-6.5) | 5.6 (4.4-6.1) | 5.8(4.3-6.7) |
| Baseline ALT level, U/L | 76±60 | 76±70 | 80±68 |

Changes in ALT level from baseline to the last visit are displayed in table 3. Nearly all patients had normalization of ALT level in Group IIa and II b. there is a significant reduction in group I, but neither of patient achieved SVR.

**Table 3:**
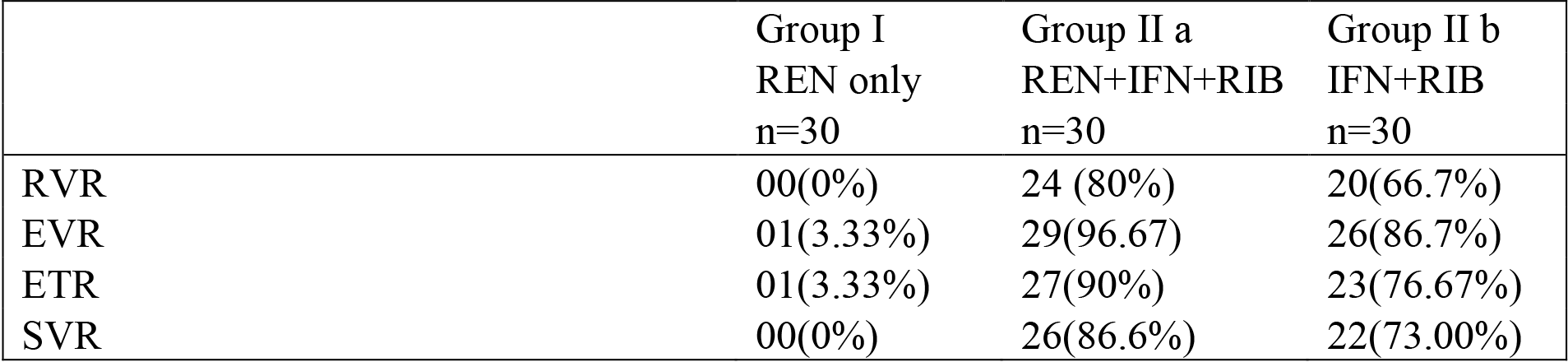
Comparison of RVR, cEVR, ETR, and SVR Rates by treatment Group Intention-To-Treat.

**Table 4:** Changes in ALT from baseline to the last visit.

|  | Group I<br>REN only<br>n=30 | Group II a<br>REN+IFN+RIB<br>n=30 | Group II b<br>IFN+RIB<br>n=30 |
| --- | --- | --- | --- |
| BASELINE ALT | 76 | 76 | 80 |
| LAST VISIT | 40 | 20 | 21 |

The virological response is observed in some patients in group I (Renessans only) but this is not statistically significant, only one patient has EVR but neither patient achieved ETR or SVR. Exploratory analyses were conducted to identify demographic or disease-related characteristics (age, sex, BMI, baseline HCV RNA, baseline ALT level) predictive of an SVR. Lower baseline HCV RNA level was an independent predictor of SVR in the standard of care group (II b) [P=0.008], but not in the other two groups. No other significant predictor of SVR were identified. Four patients could not complete the therapy due to Serious Adverse Event Two in group IIa and Two in group IIb. Seven patients in group I were lost to follow up. These patients were recorded as Non-Responders.

### ADVERSE EVENTS

The most common adverse events in group II (a&b) were anemia, leucopenia, neutropenia, and thrombocytopenia, all typical of interferon and Ribavirin therapy. Common side effects in group I was epigastric discomfort and distaste. Four adverse events were serious, two in group IIa had grade IV thrombocytopenia and acute psychosis and two in group IIb had grade IV thrombocytopenia and atypical pneumonia, all required hospitalization. 12 patients had a dose reduction of Ribavirin due to anemia, five in group II and seven in group II b.

## DISCUSSION

We concluded this study successfully and our results were quite encouraging. Patients in this study belonged to HCV genotype 3. Our results indicate that the higher SVR rates in group IIa were associated with higher RVR rates as shown in table 3. The proportion of patients achieving an RVR with triple therapy was significantly higher than for the standard of care dual therapy (80% vs. 66.7 % p=0.02) table 3. An important limitation of the study is that all the patients enrolled were having genotype 3 as this genotype is around 90 % among chronic HCV infected patients in Pakistan. The SVR rate for treatment of HCV genotype 3 patients using the standard of care (interferon) is 68%. The rationale for the Renessans study was based on an initial pilot experience that showed some antiviral activity when administered alone in patients with chronic HCV infected with genotype 3. This study demonstrated an increased SVR when Renessans has been added to the standard of care regimen (17).

In this study, triple therapy with Interferon, Ribavirin, and Renessans achieved an SVR rate of 86.6 % in treatment naïve patients with chronic HCV infected with genotype 3, which was superior to SVR achieved with the standard of care using Interferon and Ribavirin 73 % (P=0.021). There were no added side effects observed which are peculiarly associated with the use of Renessans.

This study reports the safety and efficacy of Renessans in combination with interferon and Ribavirin. Further multicentre studies required to confirm these findings. The results of this study show that Renessans has the potential to increase SVR and further studies required to test these results in non-responders and relapser patients and in those who are infected with genotype 1 and 4 which has low SVR rates with a standard of care treatment compared to genotype 3.

## Data Availability

We have all data available. Will include more data if required.

https://clinicaltrials.gov/ct2/show/NCT01463592

